# Characterising global risk profiles of Mpox clade Ib importation

**DOI:** 10.1101/2024.09.09.24313259

**Authors:** Toshiaki R. Asakura, Sung-mok Jung, Shihui Jin, Gang Hu, Akira Endo, Borame Lee Dickens

**Author notes:** Correspondence to: Akira Endo, MD, PhD, 12 Science Drive 2, #10-01, Singapore 117549.

## Abstract

The novel mpox clade Ib initially identified in the Domestic Republic of Congo has spread to its multiple neighbouring countries as well as countries beyond the African continent. We characterised the global risk of importation of mpox clade Ib, highlighting the need to ramp up surveillance capacity for early detection.

## Main text

In September 2023, a cluster of mpox cases were reported in Kamituga health zone of the South Kivu province, the Domestic Republic of Congo (DRC), from which a novel virus clade Ib was identified.^1^ Genetic analyses have shown evidence of sustained human-to-human transmission since its emergence,^1^ with rapid spread to other health zones in South Kivu and neighbouring countries, including Burundi, Rwanda, Uganda and Kenya by August 2024. Unlike clade Ia, primarily driven by zoonotic exposure and limited human-to-human transmission typically within households,^2^ clade Ib has been reported to be additionally spread through sexual contact, highlighted by the increased risk among sex workers and their clients.^3^ This shift in transmission modes and rapid spread prompted the (re-)declaration of the World Health Organization’s Public Health Emergency of International Concern (PHEIC) on mpox. Shortly after, clade Ib reached beyond the African continent with two travel-associated cases reported in Sweden and Thailand on 15 and 22 August 2024, respectively.^4^

International flight travel volume data can help to infer and contextualise the country-specific risk of importing clade Ib mpox cases into countries currently without local transmission (Figure 1A). We used a simple statistical model assuming the risk of importation in a country is proportional to the OAG flight travel volume (monthly average from May 2023 to June 2024) from the source countries. Given the two importations observed outside Africa, we estimate that 23,183 (95% confidence interval: 3,765-72,255) local clade Ib infections may have accumulated in the DRC, assuming DRC is the primary source of exportation and that no additional undetected importations exist (see supplementary materials for sensitivity analysis assuming additional source countries). As of August 2024, the reported numbers of confirmed and suspected cases in the North and South Kivu provinces—where clade Ib is known to circulate in the DRC—are 1,443 and 5,077, respectively. This discrepancy may suggest moderate underreporting of clade Ib infections in the DRC.

**Figure 1.**
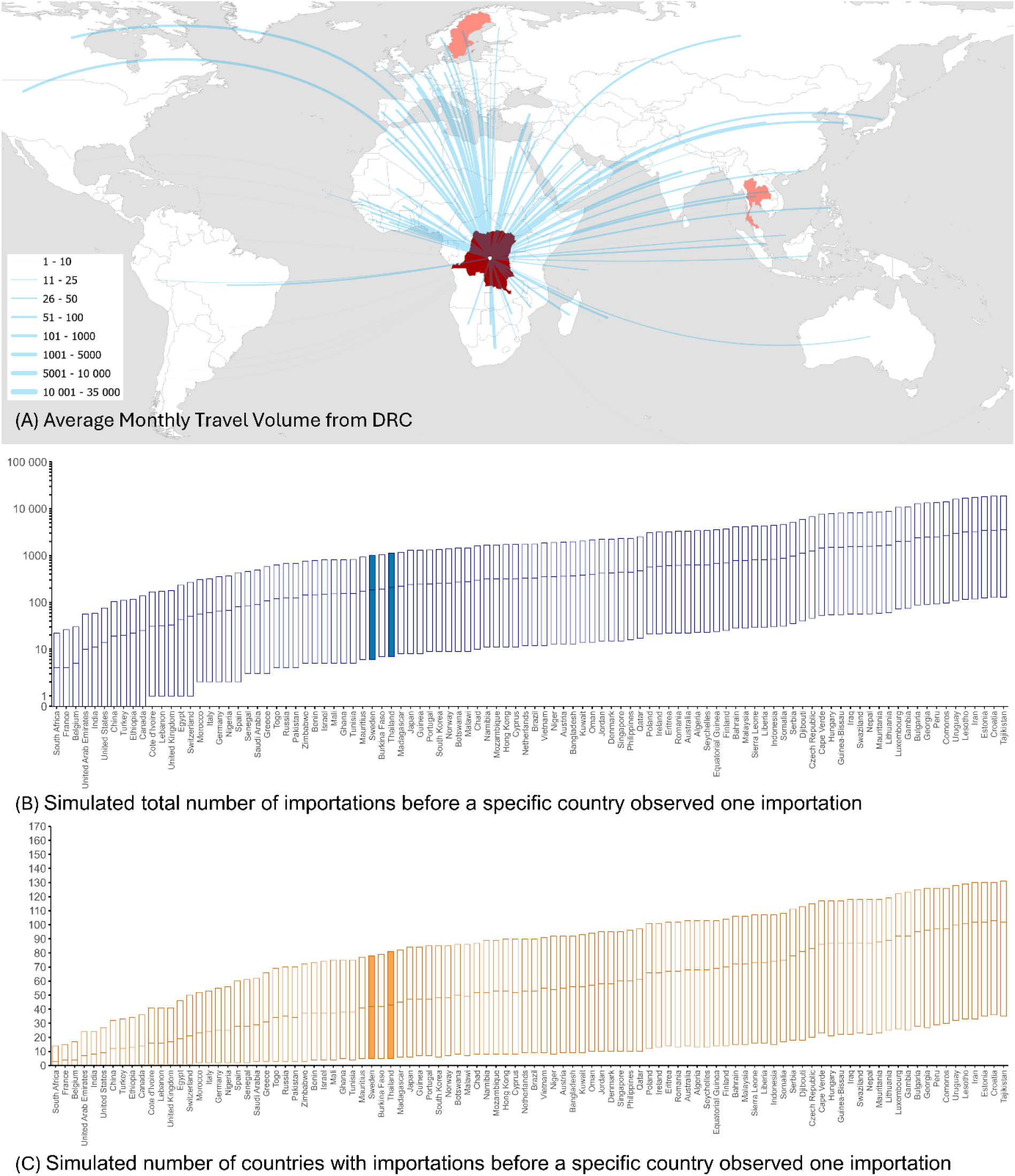
Travel volumes and simulated number of importations and countries before the first importation in a specific country. **(A)** Travel volume from the DRC to international countries. **(B)** Simulated total number of importations before a specific country observed one importation and **(C)** Simulated number of countries with importations before a specific country observed one importation. Only the top 100 countries with the highest travel volume are included in the figure. Sweden and Thailand reporting an imported clade Ib case are highlighted. This analysis assumes that DRC is the only source of exportation while sensitivity analyses assuming the additional countries as sources (including Burundi, Uganda, Kenya and Rwanda) are available in the Supplementary Materials (Figure S1, S2). Flight volumes for the above countries to Angola, Burundi, Cameroon, DRC, Central African Republic, Gabon, Kenya, Rwanda, Republic of the Congo, South Sudan, Tanzania, Uganda and Zambia were excluded as they have reported historic or current clade Ia or Ib mpox cases.

We simulated the likely phase at which a country may experience its first clade Ib importation (Figure 1B-C) by estimating how many imported cases and countries would be recorded before the first importation occurs. This provides an overview of the clade Ib importation landscape, showing a likelihood of immediate importation in a country based on importations reported elsewhere. We obtained similar results in our sensitivity analysis where we assumed countries with confirmed cases other than the DRC are also sources of exportation (Figure S1, S2).

Our simulations suggest that neither Sweden (36th [95% range: 3-74th] country to import in our simulation) nor Thailand (47th [6-88th]), reporting the first two importations, is among the countries to expect the earliest importation. Whilst the full travel history of these two cases including origin, destination and stopovers may explain this discrepancy, it also suggests potential undetected importations in countries with high travel volume. Additionally, clade Ib’s deletion in the target domain could have affected PCR testing accuracy for the clade differentiation from globally circulating clade IIb,^5^ especially before the PHEIC declaration. Enhancing surveillance capacity worldwide to detect possible clade Ib arrivals is crucial to prevent further international spread, which would also allow the global health community to sustain support for containment in the most affected region.

## Author contributions

TRA: Conception, Investigation, Formal analysis, Writing-original draft. S-mJ: Investigation, Critical review and editing of the manuscript. SJ: Data curation. GH: Data curation. AE: Conception, Investigation, Formal analysis, Supervision, Critical review and editing of the manuscript. BLD: Investigation, Formal analysis, Supervision, Critical review and editing of the manuscript.

## Data availability

The data analysis code in the present study is deposited on https://github.com/toshiakiasakura/mpox1b_importation_risk. The OAG flight volume data was not uploaded as per the terms of the contract.

## Supplementary information

## Material and methods

### Data source

We used the OAG flight volume dataset spanning from May 2023 to June 2024, which provides comprehensive information on flight volumes between individual airports worldwide.^1^ In the present study, we aggregated airport data to obtain country-specific flight volumes and took the monthly average over the included months. The flight volume between any two countries includes direct flights and indirect flights that involve transit through third countries.

Population size data for the Democratic Republic of Congo (DRC), Burundi, Rwanda, Uganda, and Kenya were retrieved from the United Nations population data for 2024.^2^

### Back-projecting the cumulative incidence in countries with ongoing spread

We estimated the cumulative incidence of mpox clade Ib in DRC at the time of importation events in Sweden and Thailand. We assumed that the probability of contracting an infection in DRC is equivalent between travellers and residents. Let *C*_t_ be the cumulative incidence of mpox clade Ib in DRC by day *t* and *O*_i,t_ be the cumulative incidence in an importing country *i* by day *t*. We model *O*_i,t_ to follow a binomial process:

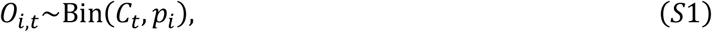

where *p*_i_ is the proportion of travellers in DRC and subsequently moving to country *i* by plane among the population of DRC (*N*_DRC_). This proportion (*p*_i_) is described by

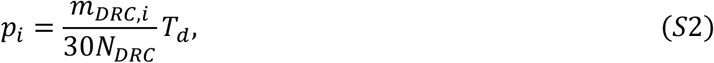

where *m*_DRC,i_ is the average monthly flight volume from DRC to country *i* and *T*_d_ is the average length of stay in DRC (assumed to be 7 or 10 days).

Even though DRC experienced mpox outbreaks across multiple provinces, the majority of mpox clade Ib were reported specifically from North or South Kivu, DRC. Despite the unavailability of specific flight volume data used by travellers visiting North/South Kivu, when we assumed that the travellers’ prevalence between North/South Kivu and other provinces within the DRC is proportional to the population size distribution, we can interpret *p*_*i*_ as if we used the flight volume and population size for North and South Kivu. We obtained the monthly flight volume from DRC to country *i* by averaging the flight volume from May 2023 to June 2024.

The likelihood function for the cumulative incidence is then described by

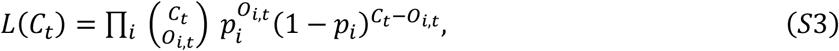

We included countries with the top 100 flight volumes from DRC, while we excluded the following countries which have reported historic or current clade I mpox cases^3^: Angola, South Suda, Tanzania, Zambia, Republic of the Congo, Central African Republic, Burundi, Rwanda, Uganda, Kenya, Cameroon, and Gabon.

Since both of the two imported cases in Sweden and Thailand visited multiple African countries, we conducted two sensitivity analyses, assuming the primary source of exportation was i) DRC and Burundi, or ii) DRC, Burundi, Rwanda, Uganda and Kenya. The combined monthly average flight volume and combined population size for those countries were used in equation S2. For example, *p*_i_ for DRC and Burundi was described by

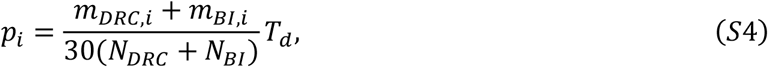

where *m*_BI,i_ is the average monthly outgoing flight volume from Burundi to country *i* and *N*_BI_ is the population size in Burundi.

We estimated *C*_t_ using the maximum likelihood estimation with the differential evolution for the optimisation algorithm. The 95% confidence interval was obtained using the profile likelihood method.

### Model for the simulated total number of importations and simulated number of countries ever importing mpox clade Ib

Assuming that the risk of importing a clade Ib mpox case from countries with ongoing spread is proportional to the flight travel volume, we simulated the cumulative number of imported cases outside most affected countries before a given country *i* experiences its first imported case: *D*_*i*_. That is, (*D*_*i*_+1)-th imported case outside of countries with an ongoing outbreak would be the first imported case in country *i*. Similarly, we also simulated the number of countries ever importing mpox clade Ib before the first importation in country *i*: *N*_*i*_. Country *i* would be the (*N*_*i*_+1)-th country to ever import a case.

Under the assumption that either (i) DRC, (ii) DRC and Burundi and (iii) DRC, Burundi, Uganda, Kenya and Rwanda are the primary source of importation throughout the time course of interest (i.e. we did not consider established local transmission in any other country onward that could contribute to importation elsewhere), we modelled *D*_*i*_ to follow a geometric distribution:

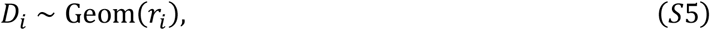

where *r*_*i*_ is the relative flight travel volume among the countries at risk of importation (all countries except the countries which have reported clade Ia or Ib). Given the cumulative number of imported cases globally before country *i* observed an importation (*D*_i_), the number of countries with nonzero importations *N*_i_ can be sampled from

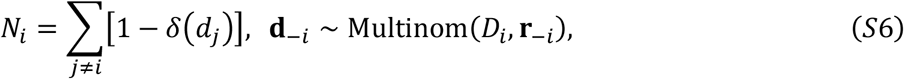

where *d*_j_ is the cumulative number of imported cases in country *j* (conditioned on *D*_i_), *δ* is the Kronecker delta, **d**_-i_ is the vector of *d*_j_ for all countries except the i-th country, and **r**_-i_ is the vector of the relative flight travel volume excluding the i-th country. We simulated the model 10,000 times for each assumption and calculated the median, 2.5h and 97.5th quantiles.

The analysis for back-projection was performed in Julia v1.7.3 and the simulation analysis was performed in R v4.1.3. The code used in the present study was deposited at https://github.com/toshiakiasakura/mpox1b_importation_risk.

## Supplementary results

### Back-projected cumulative incidence in countries with ongoing outbreaks

Based on the WHO situation report as of 29 August 2024,^4^ the reported numbers of confirmed and suspected mpox cases in the North and South Kivu provinces—where clade Ib is known to circulate in the DRC—are 1443 and 5007, respectively.

**Table S1.** Estimated cumulative incidence of mpox clade Ib for each pattern of the primary source of importation.

| Date | Cumulative reported cases | $T_d$ (days) | Estimated cumulative incidence (95% confidence interval) for the primary source of importation | | |
| --- | --- | --- | --- | --- | --- |
|  |  |  | DRC<sup>a</sup> | DRC, Burundi | DRC, Burundi, Rwanda, Uganda, Kenya |
| 2024-08-15 | Sweden (1) | 7 | 11,591<br>(634-51,624) | 11,639<br>(637-51,839) | 3,614<br>(198-16,095) |
|  |  | 10 | 8114<br>(444-36,137) | 8148<br>(446-36,287) | 2,530<br>(138-11,266) |
| 2024-08-22 | Sweden (1),<br>Thailand (1) | 7 | 23,183<br>(3,765-72,255) | 23,279<br>(3,781-72,556) | 7,227<br>(1,174-22,527) |
|  |  | 10 | 16,228<br>(2,636-50,579) | 16,295<br>(2,647-50,789) | 5,059<br>(822-15,769) |

<sup>a</sup> Estimates for DRC can also be interpreted as the cumulative incidence in North and South Kivu if we assume the travellers' prevalence between North/South Kivu and other provinces within the DRC is proportional to the population size distribution.

Sensitivity analysis for the simulated number of importations and number of countries ever importing given one observation in a specific country

**Figure S1.**
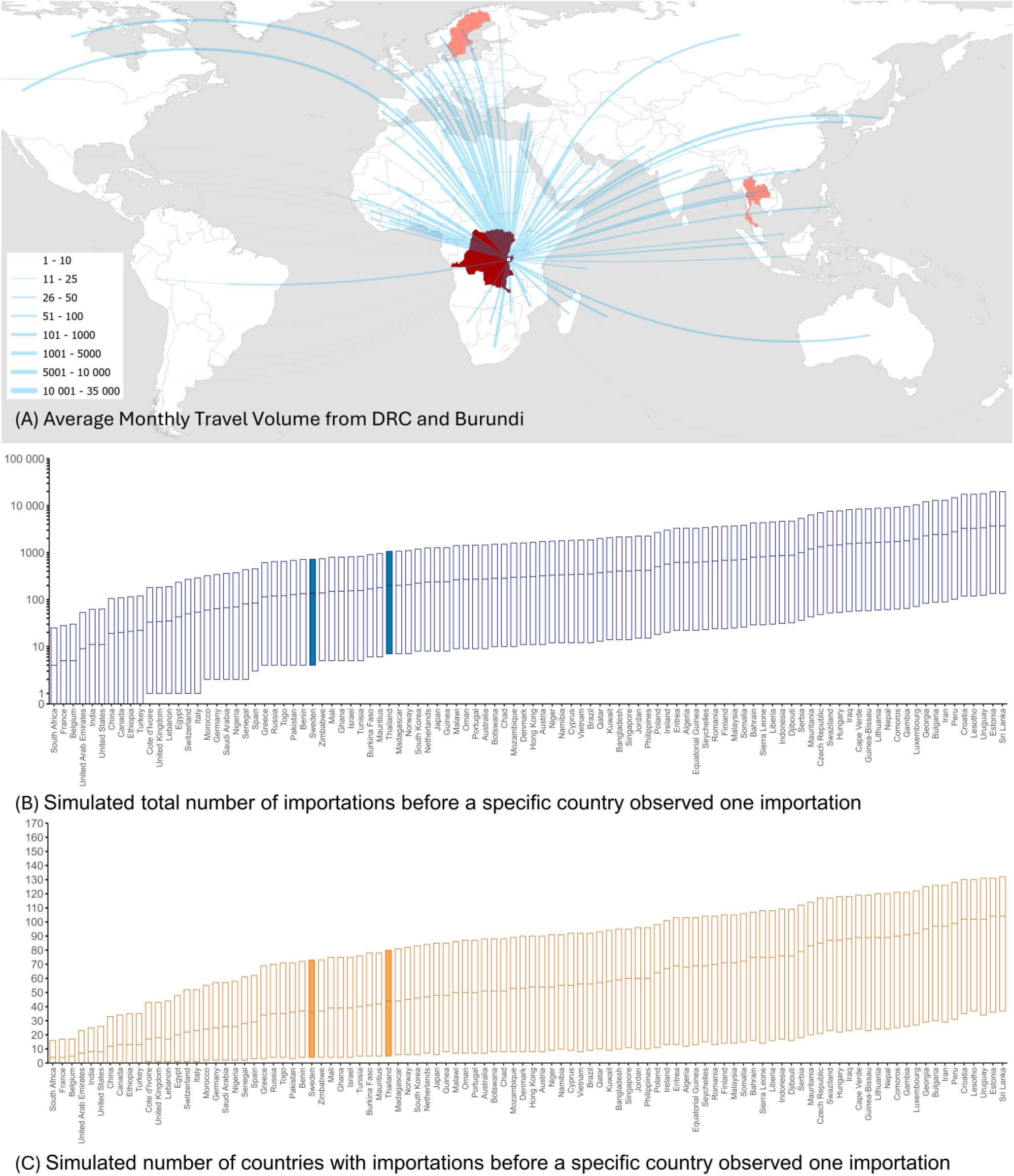
Travel volumes, and simulated number of importations and countries before the first importation in a specific country. **(A)** Travel volume from the DRC and Burundi combined to international countries. **(B)** Simulated total number of importations before a specific country observed one importation and **(C)** Simulated number of countries with importations before a specific country observed one importation. Only the top 100 countries with the highest travel volume are included in the figure. Flight volumes for the above countries to Angola, Burundi, Cameroon, DRC, Central African Republic, Gabon, Kenya, Rwanda, Republic of the Congo, South Sudan, Tanzania, Uganda and Zambia were excluded as they have reported historic or current clade Ia or Ib mpox cases.

**Figure S2.**
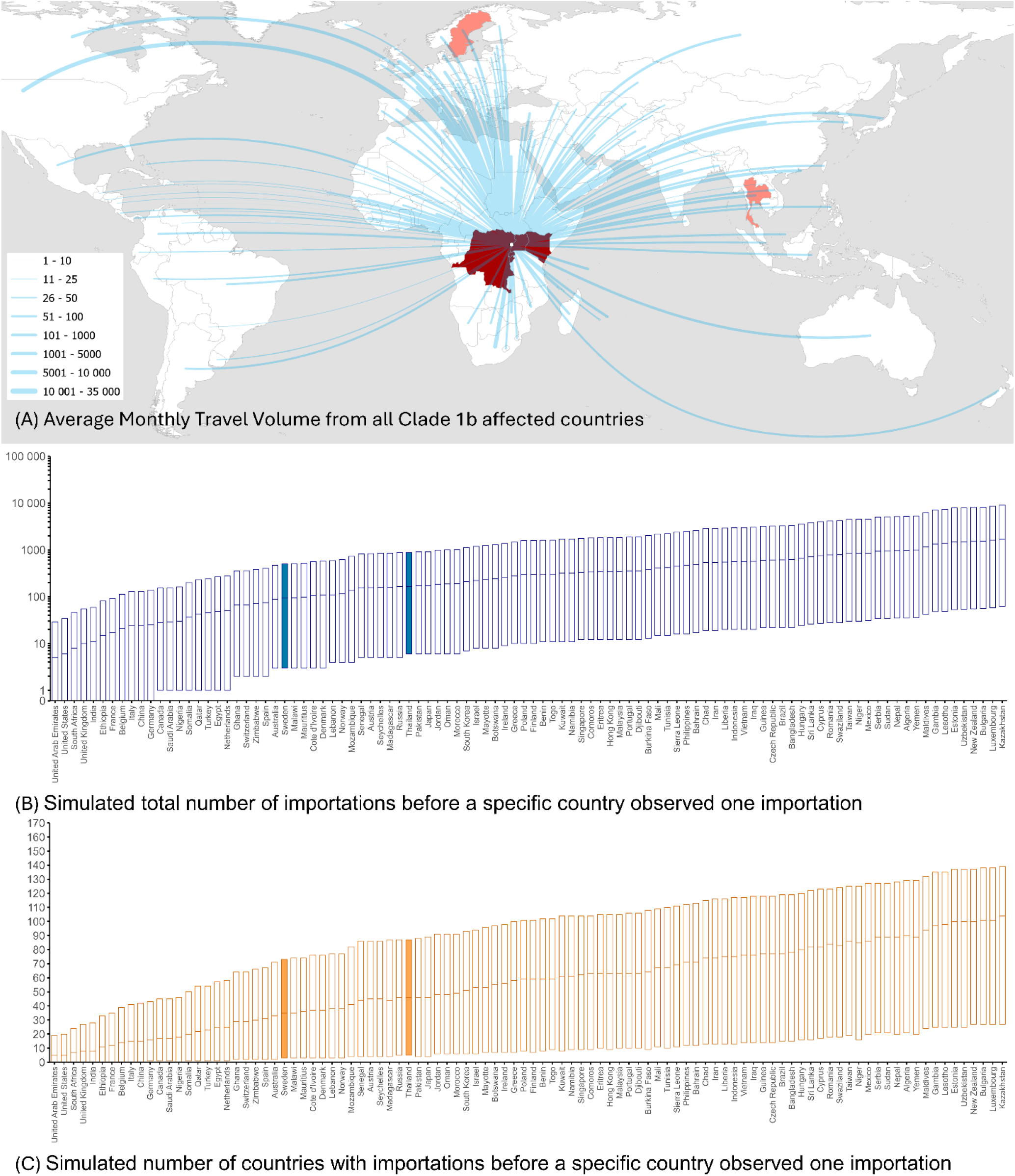
Travel volumes, and simulated number of importations and countries before the first importation in a specific country. **(A)** Travel volume from the DRC, Burundi, Uganda, Kenya and Rwanda combined to international countries. **(B)** Simulated total number of importations before a specific country observed one importation and **(C)** Simulated number of countries with importations before a specific country observed one importation. Only the top 100 countries with the highest travel volume are included in the figure. Flight volumes for the above countries to Angola, Burundi, Cameroon, DRC, Central African Republic, Gabon, Kenya, Rwanda, Republic of the Congo, South Sudan, Tanzania, Uganda and Zambia were excluded as they have reported historic or current clade Ia or Ib mpox cases.

